# Anatomically optimised structural connectome derivation for the neonatal brain

**DOI:** 10.64898/2026.10.20.26366012

**Authors:** Calvin K. Hew, Simona Leserri, Dogu Baran Aydogan, Mark E. Bastin, Alexandra F. Bonthrone, Kirsty Paterson, Gayle Barclay, Charlotte Jardine, Keith M. Smith, Michael J. Thrippleton, Riccardo Marioni, James P. Boardman, Manuel Blesa Cabez

## Abstract

Structural connectomics provides a means of characterising white matter (WM) network organisation using diffusion MRI (dMRI), but reconstructing neonatal structural connectomes remains challenging due to the immature and heterogeneous nature of the developing brain. Although recent neonatal studies have increasingly adopted approaches beyond conventional diffusion tensor imaging (DTI), there remains scope to incorporate more advanced tractography and streamline post-processing approaches within neonatal pipelines. Here, we propose a neonatal structural connectome pipeline integrating surface-based anatomical constraints, ensemble tractography, hierarchical anatomical refinement of pathways (HARP) filtering, and spherical-deconvolution informed filtering (SIFT2) reweighting. We acquired 3T MRI data from 300 neonates (185 preterm, 115 term-born) to assess the impact of pipeline configuration on network organisation. Mean gestational age (GA) at birth was 29.31 (23.43–32.86) weeks in preterm infants and 39.52 (37.00–42.43) weeks in term infants. At scan, mean GA was 40.85 (36.29–49.00) weeks in preterm infants and 42.06 (38.29–48.00) weeks in term infants. Four connectome variants were generated: raw surfaced-based tractograms, HARP-filtered, SIFT2-weighted, and combined HARP+SIFT2. Pipeline performance was primarily evaluated using an existing brain age prediction model: connectome-based predictive model (CPM) and our derived connectomes to estimate GA at scan in term and preterm infants. In addition, global graph theoretical metric analyses were conducted to provide a complementary assessment of pipeline configuration differences in network organisation. In total, seven graph theoretical metrics were calculated and evaluated: global efficiency, local efficiency, transitivity, modularity, assortativity, degree variance and statistical complexity. Across all configurations, connectomes supported accurate brain age prediction, with mean absolute errors (MAE) of 0.76–0.79 weeks in term infants and 1.00–1.06 weeks in preterm infants. Prediction error and brain age gap variability were greater in preterm than term infants across configurations, with HARP+SIFT2 achieving the lowest preterm MAE (1.00 weeks) and brain age gap (BAG) variability (SD = 1.38 weeks). However, differences in prediction performance across configurations were minimal, suggesting limited influence of pipeline choice on brain age prediction. However, analysis of global network metrics demonstrated that pipeline choice significantly influenced downstream measures of network organisation, with HARP+SIFT2 being the only configuration to identify significant preterm vs. term differences across all seven metrics examined. These findings demonstrate the importance of considering tractography and streamline-processing choices when constructing neonatal structural connectomes and provide a framework for more anatomically informed investigation of early brain development.

**Issue Section:** Research

## 1. Introduction

Traditional approaches for modelling neonatal white matter connectivity often fail to capture the full complexity of developing networks, limiting the biological plausibility of derived structural networks (Farquharson et al., 2013). Advanced network construction methods improve the anatomical plausibility of reconstructed streamlines and connectomes. These include the use of sophisticated tractography algorithms (Aydogan & Shi, 2021), anatomically-constrained tractography (ACT) (Smith et al., 2012), surface-based techniques (Yeh et al., 2017), streamline filtering approaches (Leserri et al., 2026) and microstructure informed filtering methods (Leserri et al., 2026; Smith et al., 2015). However, the application of these advanced techniques in neonatal populations remains limited (Verschuur et al., 2025) due to additional methodological and technical challenges unique to this population (Verschuur et al., 2024). For example, increased partial volume effects (Verschuur et al., 2025), immature white matter (Berman et al., 2005) and reduced image contrast (van Pul et al., 2006) complicate fibre orientation estimation and network analysis.

**Figure 1.**
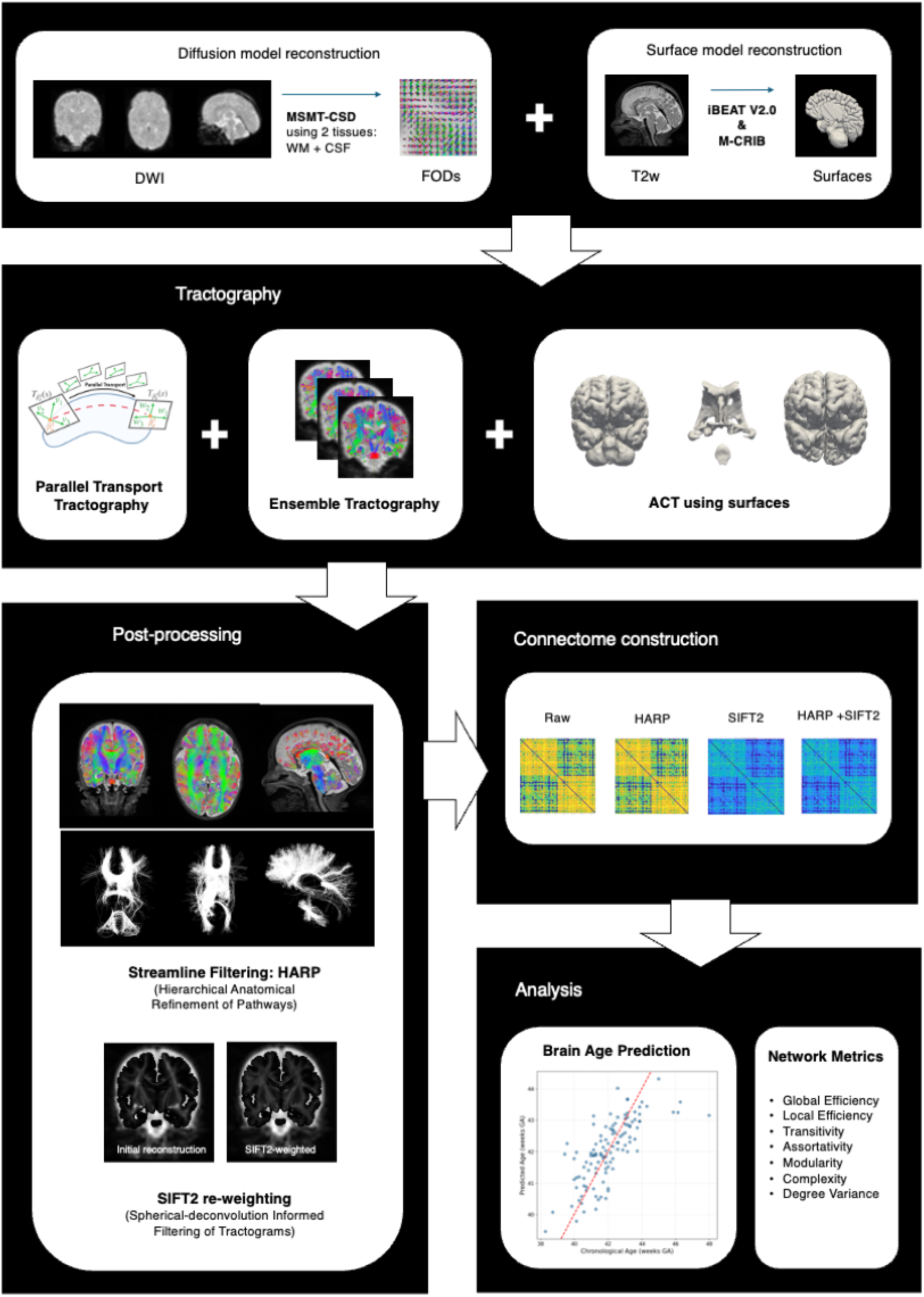
Overview of processing pipeline and analysis

Preterm birth is a major risk factor for long term neurodevelopmental impairments, including cognitive deficits, behavioural challenges, and atypical brain network development (Batalle et al., 2017; Johnson & Marlow, 2017). Preterm vs. term comparisons offer a clinically relevant example through which the effect of different connectome reconstruction approaches can be examined. As such, methodological choices including streamline filtering such as Hierarchical Anatomical Refinement of Pathways (HARP) (Leserri et al., 2026; Leserri & Aydogan, 2024) and reweighting strategies such as Spherical-Deconvolution Informed Filtering of Tractograms (SIFT2) (Smith et al., 2015), are reported to affect the sensitivity of connectome-derived metrics to group-level differences (Frigo et al., 2020; McColgan et al., 2018).

Structural brain dysconnectivity can be studied using diffusion MRI (dMRI) tractography, providing valuable insight into the impact of preterm birth on white matter maturation and large-scale brain networks (Blesa et al., 2021; Galdi et al., 2020; Wheater et al., 2022). The limitations of Diffusion Tensor Imaging (DTI) for dMRI analysis are well established, particularly its inability to resolve complex fibre orientations which motivated the development of more advanced diffusion reconstruction methods to model multiple fibre orientations (Schilling et al., 2018). However, the current state of the art in neonatal diffusion analysis remains predominantly reliant on DTI-based tractography, with more advanced approaches such as constrained spherical deconvolution (CSD) yet to be widely adopted, according to an extensive review by Vershuur et. al. (2025). Recent improvements in neonatal diffusion MRI acquisition and the release of publicly available high-quality neonatal datasets (Boardman et al., 2020, 2024; Edwards et al., 2022; Howell et al., 2019) now provide an opportunity to adopt more sophisticated tractography frameworks that address these limitations. For example, some groups have started to adopt more advanced structural connectome pipelines in neonates combining ACT with some microstructure informed filtering methods (Batalle et al., 2017; Blesa et al., 2019, 2021; Sa de Almeida et al., 2021; Taoudi-Benchekroun et al., 2022). Thus, we identify an important need to further evaluate the applicability of advanced diffusion MRI modelling techniques for analysing structural brain development in the neonatal period. In this work, we aim to build upon these methodological advances by combining multi-tissue constrained spherical deconvolution (MSMT-CSD) with surface-based anatomical constraints and microstructure informed streamline filtering to improve the anatomical plausibility of neonatal structural connectomes.

Structural connectomes can be derived from tractography reconstructions (Wu et al., 2026) and evaluated using machine learning approaches that assess their ability to capture age related patterns of brain development (Brown & Hamarneh, 2016). Brain age prediction has emerged as a useful approach for evaluating developmental information within neonatal connectomes, with models trained to predict an individual’s postmenstrual or gestational age from connectivity features (Brown et al., 2017; Kawahara et al., 2017; Sun et al., 2024; Taoudi-Benchekroun et al., 2022). In addition, connectomes are often used to conduct graph-theoretical network analysis, allowing quantification of topological properties such as integration (Latora & Marchiori, 2001), segregation (Newman, 2003, 2006), organisation (Leung & Chau, 2007) and complexity (Yeung et al., 2025).

This study aims to provide a framework for neonatal structural connectome construction, building upon advancements beyond conventional DTI-based approaches through the integration of surface-based anatomical constraints for tractography, ensemble tractography and the additional of HARP-based streamline filtering in conjunction with SIFT2 reweighting. To evaluate our pipeline, we will examine the value of four post-processing configurations: unfiltered, HARP-filtered, SIFT2-weighted and a combination of HARP+SIFT2. Connectomes derived from these pipelines will be evaluated using a brain age prediction model (Sun et al., 2024) and through global network metrics analysis.

## 2. Methods

### 2.1. Dataset

A total of 300 neonates were included in the study, comprising 185 preterm (born at <33 weeks) and 115 term infants (born > 37 weeks). Families were recruited from the Royal Infirmary of Edinburgh (RIE), NHS Lothian, and underwent MRI at the Edinburgh Imaging Facility RIE, University of Edinburgh (Boardman et al., 2020, 2024), following written informed parental consent. The study has ethics approval from the South-East Scotland Research Ethics Committee 02 (16/SS/0154 and 23/SS/0067) and NHS Lothian Research and Development (16/SS/0154 and 2023/0150). Preterm infants were eligible if their mothers delivered at the study centre or were transferred to the study centre in utero because of anticipated delivery before 33 weeks of gestation. Preterm infants were excluded from the study if they: (1) were transferred to the study centre after birth for intensive care, (2) had congenital anomalies (defined as structural/functional abnormalities arising during intrauterine life) detected before birth, at birth, or later in life), or (3) had contraindications to undergoing 3T MRI according to the Edinburgh Imaging Facility safety policy and UK Medicines and Healthcare Products Regulatory Agency guidelines. Participant characteristics are shown in Table 1.

**Table 1.** Characteristics of term and preterm participants including sex (M:F), gestational age at birth (weeks), gestational age at scan (weeks), birth weight (kg) and scan weight (kg).

|  | Term (N=115) | Preterm (N=185) | Preterm vs Term |
| --- | --- | --- | --- |
| Sex (Male : Female) | 50.4% : 49.6% | 58.4% : 41.6% | $p = 0.18$ |
| GA at birth (weeks) | 39.52 (37.00–42.43) | 29.31 (23.43–32.86) | $p < 0.001$ |
| GA at scan (weeks) | 42.06 (38.29–48.00) | 40.85 (36.29–49.00) | $p < 0.001$ |
| Birth weight (kg) | 3.43 (2.41–4.56) | 1.28 (0.45–2.38) | $p < 0.001$ |
| Scan weight (kg) | 3.62 (2.60–4.93) | 3.05 (1.56–5.28) | $p < 0.001$ |

Participants were scanned using a clinical 3T Siemens MAGNETOM Prisma scanner (Siemens Healthcare, Erlangen, Germany) with a 16-channel paediatric head coil. Structural images were acquired using a 3D T2-weighted SPACE sequence with 1-mm isotropic resolution. Diffusion-weighted MRI data were collected using an axial whole-brain multi-shell acquisition comprising 3 diffusion weighted directions at b = 200 s/mm^2^, 6 directions at b = 500 s/mm^2^, 64 directions at b = 750 s/mm^2^, and 64 directions at b = 2500 s/mm^2^, alongside 19 b = 0 s/mm^2^ volumes (including 3 reverse phase-encoded volumes). An optimal angular coverage for the sampling scheme was applied (Caruyer et al., 2013). All dMRI volumes were acquired using single-shot spin-echo echo planar imaging (EPI) with 2-fold simultaneous multislice, 2-fold in-plane parallel imaging acceleration, and 2-mm isotropic voxels, with TR/TE = 3400/78.0 ms. Volumes affected by motion artifacts were re-acquired as required; dMRI acquisitions were repeated if signal loss was seen in 3 or more volumes. Infants were fed and wrapped and allowed to sleep naturally in the scanner. Pulse oximetry, electrocardiography, and temperature were monitored. Flexible earplugs and neonatal earmuffs (MiniMuffs, Natus) were used for acoustic protection. All scans were supervised by a doctor or nurse trained in neonatal resuscitation.

### 2.2. Processing Pipeline

#### Diffusion MRI Preprocessing

Preprocessing of the dMRI data included the following steps: denoising (Veraart et al., 2016) to improve the signal-to-noise ratio, Gibbs ringing removal to remove ringing artifacts (Kellner et al., 2016), eddy current, motion and EPI geometric distortion correction with outlier replacement, susceptibility-by-movement correction and slice-to-volume motion (Andersson et al., 2003, 2016, 2016, 2017, 2018, 2018; Andersson & Sotiropoulos, 2016; Smith et al., 2004) and bias field correction (Tustison et al., 2010).

#### Surface Generation

This study generates high quality anatomical surfaces to define tissue boundaries and inform streamline placement during tractography. These surfaces are incorporated into an anatomically constrained tractography (ACT) framework, which uses the reconstructed anatomy to determine where streamlines can originate, terminate, and propagate (Smith et al., 2012).

Conventional ACT typically relies on voxel-based anatomical masks, which may provide less precise delineation of tissue boundaries in the neonatal brain, where tissue contrast and anatomical boundaries are less clear (van Pul et al., 2006). While not specifically developed for tractography, previous studies have demonstrated that surface-based approaches offer significant advantages over volumetric approaches when applied to neonatal data (Robinson et al., 2018; Williams et al., 2023), particularly in preserving cortical topology and improving between-subject alignment. Moreover, surface-based approaches have been applied to tractography in the adult brain, with surface informed seeding and anatomical constraints showing improvements in the biological plausibility of reconstructed streamlines compared to volumetric masks (Leserri & Aydogan, 2024; Yeh et al., 2017).

Cortical white matter (WM) and grey matter (GM) surfaces were generated using iBEAT v2 (Wang et al., 2023), a multisite applicable deep learning based pipeline for neonatal cortical surface generation. This pipeline produces white matter and pial cortical surfaces derived from T2-weighted images. iBEAT v2 was used for cortical surface reconstruction due to its robust handling of neonatal brain anatomy and its ability to generate well-aligned WM/GM surfaces (L. Wang et al., 2023). In the case of all non-cortical structures, including cerebral spinal fluid (CSF), subcortical grey matter (sGM), brainstem and cerebellum, surfaces were generated directly from volumetric segmentations. Specifically, anatomical labels derived from the M-CRIB (Alexander et al., 2017) parcellation were converted into surface meshes using a label-to-surface reconstruction approach using tools from nibrary: neuroimaging library package (Aydogan, 2026).

#### Tractography

Tractography was performed on fibre orientation distributions (FODs) estimated using multi-shell multi-tissue constrained spherical deconvolution (MSMT-CSD) (Jeurissen et al., 2014). FODs were calculated using a two tissue MSMT-CSD model comprising WM and CSF as GM and WM water diffusion signals significantly overlap in the neonatal brain (Pietsch et al., 2019). Average WM and CSF response functions were estimated from a representative subset of pre-processed participants including both term-born and preterm infants. Tractography was then performed through the software Trekker (Aydogan, 2025) with the probabilistic parallel transport tractography (PTT) algorithm (Aydogan & Shi, 2021). PTT implements a probe-based tracking approach in which probes are used to model cylindrical fibre bundles and multiple candidates are checked to determine the fibre path (Aydogan & Shi, 2021). Each candidate probe is scored and evaluated for how well it aligns with the local FOD data, and the final propagation path is determined in a probabilistic manner rather than by a deterministic approach. This allows the tractography to better navigate regions with complex fibre configurations, improving the anatomical plausibility of streamlines.

Trekker’s (Aydogan, 2025) framework was used to implement ACT (Smith et al., 2012) through the incorporation of cortical and subcortical surface mesh models. To accommodate the unique anatomical characteristics of the neonatal brain and the consequent surface reconstructions, Trekker was modified to support these mesh-based constraints (see Code Availability for the trekker_neonatal fork). Streamlines were seeded from a combined surface comprising WM, brainstem and cerebellum. Trekker enforces anatomical constraints on the reconstructed tractograms through the following pathway rules:

1. *Required to be intracerebral*: streamlines were required to terminate inside GM, brainstem or cerebellum to ensure biologically plausible endpoints.
2. *Discard:* streamlines entering CSF were discarded to avoid projections into non-axonal tissue.
3. *Stop before exit*: streamlines were terminated if they reached the boundary of cortical GM.
4. *Stop before entry*: streamlines were terminated at the boundary of the brain mask to prevent trajectories from extending outside the brain.
5. *Allow traversal of sGM*: streamlines could terminate inside or traverse through subcortical regions.

Furthermore, tractography was performed using an ensemble framework (Takemura et al., 2016). As no single set of tracking parameters is considered optimal for neonatal tractography, 10 million streamlines were generated per subject by combining eight tractograms reconstructed using distinct tracking parameter settings. These settings included carefully selected values for probe length, probe radius and minimum radius of curvature, which collectively expand the local search space during streamline propagation. The eight parameter configurations are detailed in Table 2. By combining tractograms generated with multiple parameter settings, ensemble tractography reduces the bias associated with any individual configuration and improves the robustness of tract reconstruction. However, this approach incurs greater computational cost, requiring approximately 10 times more computation time than a tractogram fully generated using default parameters.

**Table 2.** Ensemble tractography parameter sets used to generate eight tractograms (1.25 million streamlines each) and their corresponding runtimes.

| Parameter set | mRC (mm) | pL (mm) | pR (mm) | mDS (mm) | sGM rule | Runtime (s) |
| --- | --- | --- | --- | --- | --- | --- |
| Tractogram 1 | 1.00e-01 | 1.00e-01 | 1.00e-01 | 1.00e-04 | ✗ | 6600 |
| Tractogram 2 | 1.00e-01 | 1.00e-01 | 1.00e-01 | 1.00e-04 | ✓ | 5809 |
| Tractogram 3 | 2.50e-01 | 2.50e-01 | 2.50e-01 | 1.00e-04 | ✗ | 4499 |
| Tractogram 4 | 2.50e-01 | 2.50e-01 | 2.50e-01 | 1.00e-04 | ✓ | 4136 |
| Tractogram 5 | 7.50e-01 | 3.75e-01 | 0 | 1.00e-04 | ✗ | 2216 |
| Tractogram 6 | 7.50e-01 | 3.75e-01 | 0 | 1.00e-04 | ✓ | 1674 |
| Tractogram 7 | 7.50e-01 | 3.75e-01 | 0 | 5.00e-02 | ✗ | 402 |
| Tractogram 8 | 7.50e-01 | 3.75e-01 | 0 | 5.00e-02 | ✓ | 303 |

*Note:*
mRC = minimum radius of curvature; pL = probe length; pR = probe radius; mDS = minimum data support.
Default values: mRC = 0.75 mm, pL = 0.375 mm, and pR = 0 mm, calculated from the smallest FOD voxel dimension (1.5 mm).
✓ = stop-before-exit rule applied using sGM; ✗ = rule not applied.
Each parameter set contributed 1.25M streamlines, resulting in a total of 10M streamlines across the ensemble.

Compared to using default settings, these parameters substantially improve tract construction in neonatal data, where FODs are less well defined. However, this comes at the cost of increased computational time and resource demands which limits practical application. An ensemble approach whereby tractograms generated using optimised parameter settings are combined with those produced using default settings, enabling broader coverage of the parameter space in a computationally feasible time frame.

#### Post processing

Probabilistic algorithms are shown to produce a large amount of spurious false positive connections in derived tractograms (Maier-Hein et al., 2017; Sotiropoulos & Zalesky, 2019). As a result, HARP (Leserri et al., 2026) and SIFT2 (Smith et al., 2015) were employed to improve the biological plausibility of the derived connectome. HARP builds on the ACT framework by enforcing layered anatomical rules to improve the plausibility of streamlines. Unlike standard ACT, HARP subdivides the brain into more granular regions. For example, separating the left and right hemispheres for WM and subcortical structures, and individually segmenting the CSF, cerebellum, and brainstem. Streamlines are then checked against these refined anatomical rules to identify and remove implausible trajectories that cross between incompatible regions, a source of false positives often overlooked in standard ACT.

SIFT2 is a technique which assigns weights to individual streamlines by comparing their densities to the FODs derived from the dMRI signal. As tractography-derived streamline counts are not quantitative measures of connectivity and implicitly assume equal contributions from all streamlines, re-weighting is necessary to reflect the actual biological strength of WM connections. Whilst HARP was used to flag and remove implausible streamlines, SIFT2 retains all streamlines instead of deleting connections to ensure internal modelling consistency between tractogram generation and filtering. By relying on the same underlying diffusion model for both tractography and filtering steps, SIFT2 avoids cross-model discrepancies, improving the reliability of the resulting connectomes.

HARP and SIFT2 were applied both independently and sequentially, resulting in four connectome variants for each participant: (i) an unfiltered (raw) connectome, (ii). HARP-filtered connectomes, (iii) SIFT2-weighted connectomes, and (iv) connectomes processed using HARP followed by SIFT2.

#### Connectome Construction

Structural connectomes were constructed by mapping whole-brain tractography to a cortical and subcortical parcellation comprising 84 regions of interest (ROI), resulting in 84×84 connectivity matrices. These ROIs were generated using the M-CRIB multi-atlas framework (Alexander et al., 2017), with labels propagated via affine registration and symmetric normalisation (SyN) (Avants et al., 2008) and merged using joint label fusion (H. Wang et al., 2013).

Weighted connectivity matrices were constructed for subsequent network metrics analysis. To further reduce potentially spurious connections in the densely connected probabilistic tractography-derived networks, we applied consistency-based thresholding, adapted from Roberts et. al. (2017). Edges were ranked according to the consistency of their connection weights across subjects, with the 30% most consistent edges retained to generate a consensus mask. A separate consensus mask was generated for each connectome type (raw, HARP, SIFT2, and HARP+SIFT2) and subsequently applied to the corresponding connectivity matrix for each subject before conducting network metric analysis.

### 2.3. Brain Age Prediction

To evaluate the ability of the constructed connectomes to predict gestational age, we applied a previously published neonatal brain age prediction model (Sun et al., 2024). This model was based on the connectome-based predictive model (CPM) (Shen et al., 2017) which estimates GA at scan from structural connectome features and has been shown to capture typical developmental trajectories of neonatal brain networks. The original study by Sun et al. (2024) tested their model using neonatal data from the Developing Human Connectome Project (dHCP) (Edwards et al., 2022). We applied this CPM model to connectomes generated from our neonatal data to assess whether the resulting networks produced accurate age predictions. Following the study design of Sun et al. (2024), predictions were performed separately for preterm and term groups to account for group-specific developmental trajectories. Prediction performance for each connectome processing approach and participant group was evaluated using mean absolute error (MAE), coefficient of determination (R^2^), correlation coefficient (r) and corresponding significance (*p*-value). Following this, the brain age gap (BAG) was calculated as the difference between predicted and observed age for each infant. BAG was used as an additional measure of developmental deviation, reflecting whether derived connectomes exhibited characteristics associated with a brain age above or below their actual gestational age.

### 2.4. Network Metrics

To characterise the topological organisation of the structural networks, a set of network metrics was computed using the Brain Connectivity Toolbox (BCT) library (Rubinovc& Sporns, 2010) in Matlab. The metrics included global and local efficiency (Latora & Marchiori, 2001), transitivity (Newman, 2003), modularity (Newman, 2006), assortativity (Leung & Chau, 2007), degree variance (Smith & Escudero, 2020) and statistical complexity (Yeung et al., 2025). These metrics capture complementary aspects of network organisation, including network integration, segregation, resilience and hierarchical organisation.

Network metrics were first computed across four connectome construction pipelines (raw, SIFT2, HARP, and HARP+SIFT2) to evaluate the extent to which different tractography filtering approaches influence the resulting network topology. As the subjects processed across all four pipelines were consistent, differences between pipeline configurations were assessed using paired *t*-tests for each pairwise comparison.

Network metrics were subsequently compared between term and preterm groups separately within each pipeline. Group differences were assessed using two-sample Welch’s *t*-tests which do not assume equal variance between the term and preterm groups. For each pipeline, false discovery rate (FDR) correction using the Benjamini-Hochberg method was applied to *p*-values obtained from the term-preterm comparisons.

## 3. Results

### 3.1. Participant characteristics

Gestational age (GA) at birth and at scanning for the 300 subjects included in this study are summarised in Table 1. GA at birth differed significantly between preterm and term groups, and a significant difference was also observed in GA at scan between the two groups (p < 0.001). The range of GA at scan was slightly wider in preterm infants (approximately 36-49) than term infants (approximately 38 - 48 weeks).

### 3.2. Tractography

Tractography was performed for each participant using eight different parameter sets and anatomical constraints derived from subject-specific surface models. Figure 1 illustrates the surfaces generated for a representative participant. The cortical GM surfaces are initially closed and therefore unsuitable for tractography, as streamline propagation cannot enter or exit areas of this region. As a result, GM surfaces were opened along the interhemispheric boundary using Euclidean distance-based criterion derived from the cortical ribbon and surrounding tissues as depicted by the left and right inner cortical GM in the figure. Incorporating surface-based seeding and ACT to our tractography enhanced the biological plausibility of streamline trajectories compared to conventional volumetric approaches.

**Figure 1.**
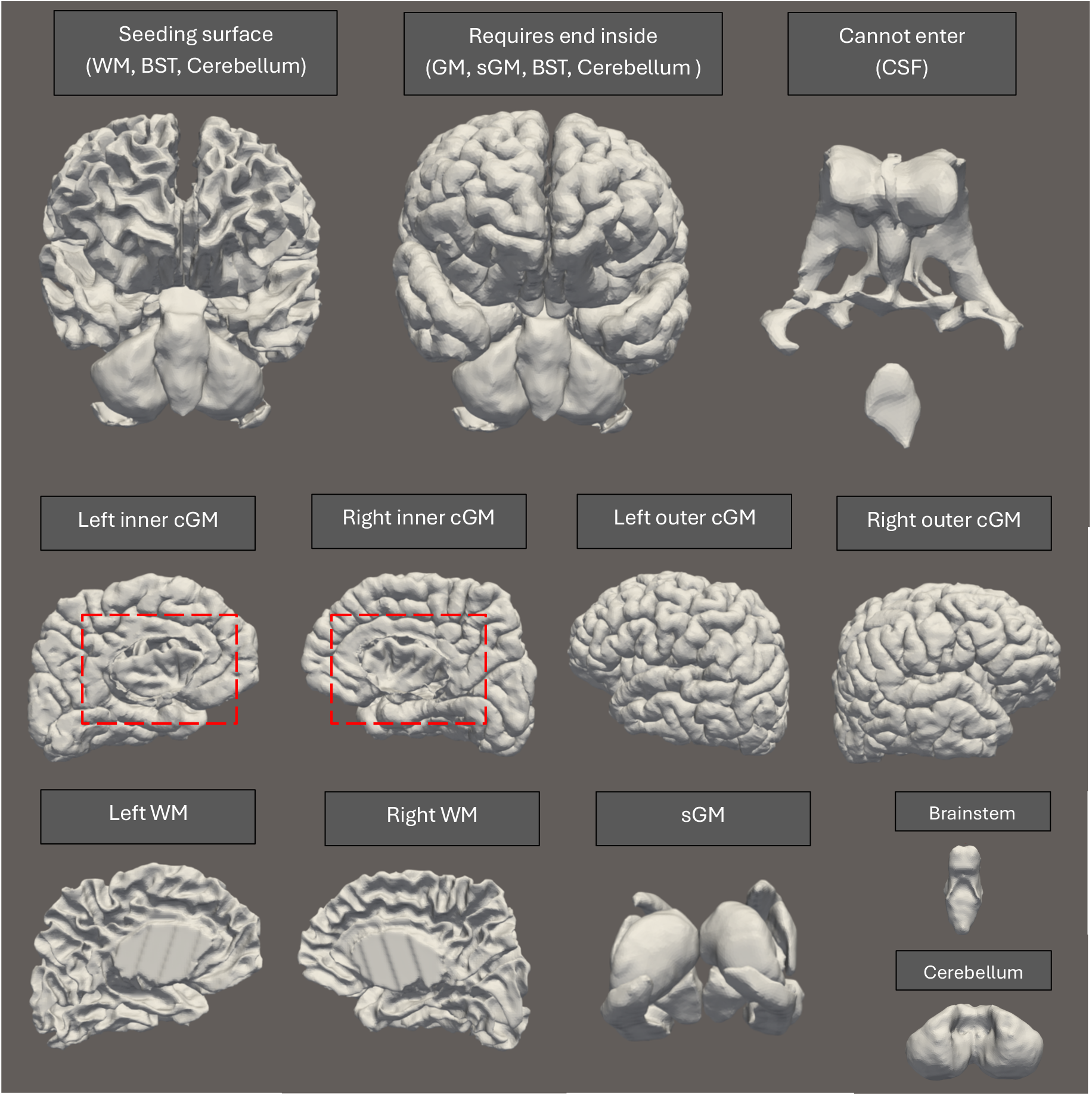
High resolution surface models used for pathway rules and tractography seeding in a single participant. Top: Combined surfaces for seeding, streamline termination, and exclusion regions. Middle: Cortical GM surfaces with cropped openings (depicted by dashed rectangles). Bottom: Remaining WM, sGM, brainstem, and cerebellum surfaces.

Table 2 summarizes the parameter sets and corresponding runtimes used to generate the ensemble tractograms. For a single participant, the eight tractograms collectively required an average processing time of 25,639 s (7.1 hours) and a peak memory requirement of approximately 4 GB RAM. Tractograms 1 - 4 (Table 2) used optimised values for minimum radius of curvature (mRC), probe length (pL), and probe radius (pR) to better model curvatures in WM tracks.

Although optimised parameter settings resulted in anatomically plausible curved trajectories during tract reconstruction (Figure 2), they incurred a substantial computational cost and required 4,136 to 6,600 s to generate 1.25 million streamlines. Extrapolating these results suggests that generating 10 million streamlines using these parameters would be impractical as it would require approximately 9 to 15 hours per participant. In contrast, tractograms generated using the default probe parameters (Tractograms 5 - 8) required approximately 300 to 2,200 seconds per subject. Default tracking parameters were calculated from the smallest FOD voxel dimension (1.5 mm), yielding mRC = 0.75 mm (½ x 1.5 mm), pL = 0.375 mm (¼ x 1.5 mm) and pR = 0. mm.

**Figure 2.**
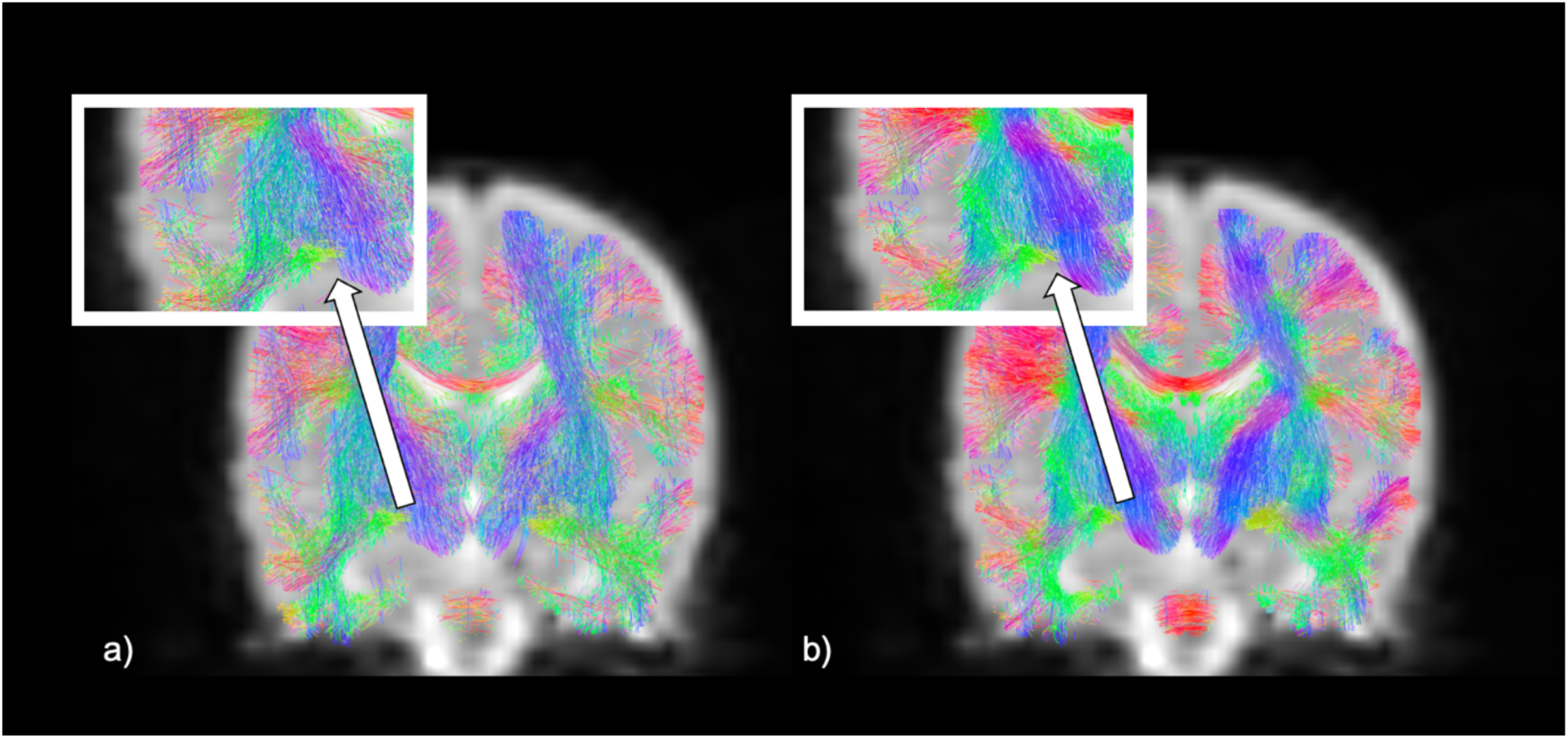
Streamline comparison between a tractogram generated using a) default parameters (Tractogram 2) and one generated with b) optimised parameters (Tractogram 8). Streamlines are colour coded by principal fibre orientation (red: left-right, green: anterior-posterior and blue: superior-inferior)

Furthermore, enforcing a minimum data support threshold (mDS = 0.0001; Tractograms 1 - 6) also increased computational time relative to tractograms generated using the default data support value of 0.05 (Tractograms 7 - 8). Whole-brain tractograms were successfully generated for each participant using an ensemble approach, yielding 10 million streamlines per connectome.

Following tractography, a further streamline filtering technique, HARP, was applied to remove anatomically implausible streamlines based on predefined neuroanatomical rules. This process selectively filtered trajectories that violated commonly overlooked anatomical constraints, such as implausible looping paths Figure 3 shows an example of streamlines removed as anatomically implausible by HARP, representing 0.09% of the 10 million generated streamlines. The small proportion of streamlines removed in this study is anticipated given the tractograms had already undergone substantial anatomical refinements through application of ACT.

**Figure 3.**
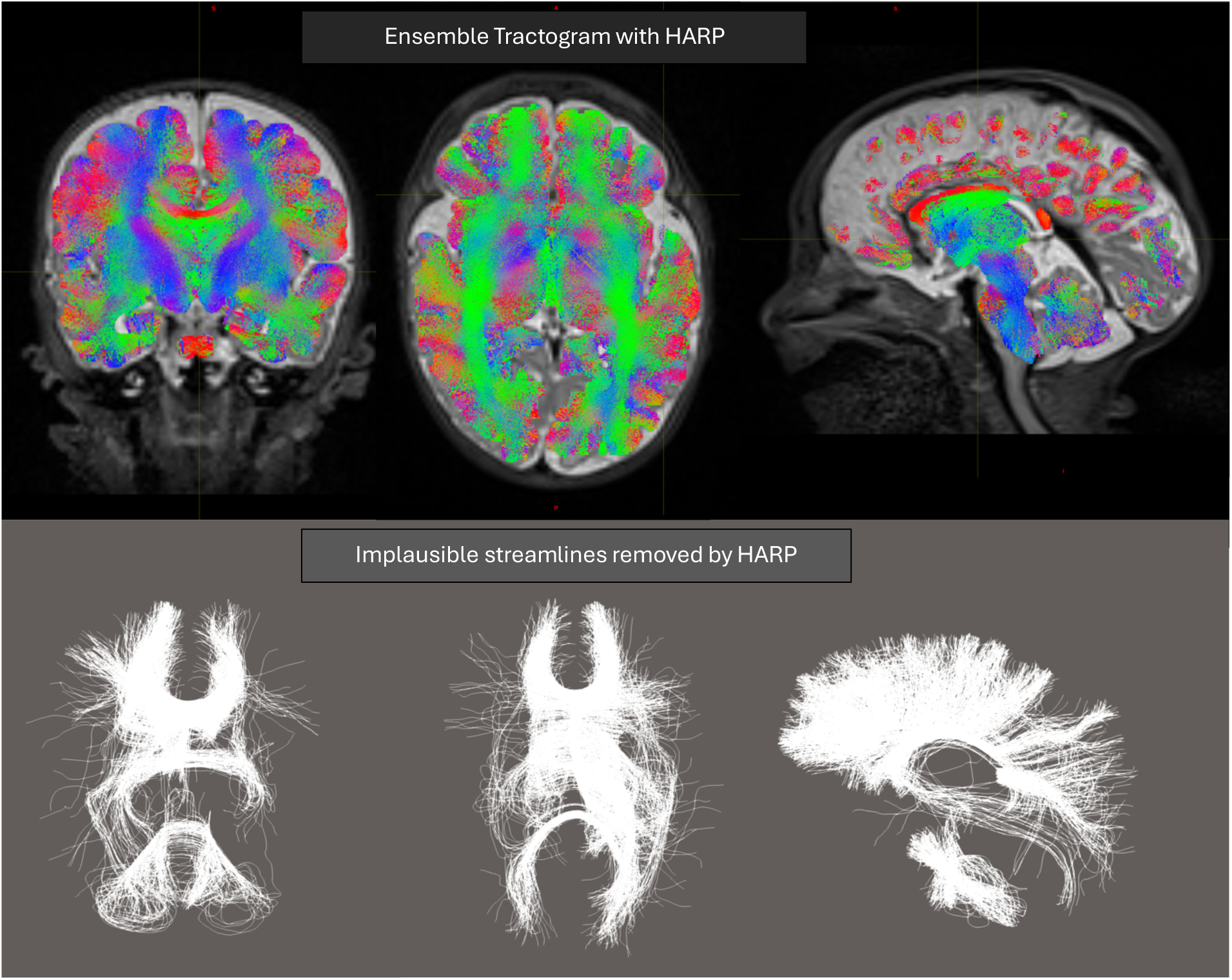
Top: Ensemble tractogram for a single participant, generated by combining tractography results from eight distinct parameter sets (each containing 1.25 million streamlines; total of 10 million) and subsequently subsampled to 500,000 for visualisation. Bottom: Implausible loop streamlines from the same participant that were identified and removed through HARP filtering (a total of 5,937 out of 10 million streamlines excluded).

### 3.3. Connectome Analysis: Age Prediction

GA in term infants were predicted with consistently high correlations and low prediction errors (Table 3). Across pipelines, prediction accuracy was greater in term than preterm infants, with MAE approximately 0.25 weeks lower in the term group. However, the differences in MAE within each group are clinically insignificant (<1 day) with only modest differences in correlation.

**Table 3:** Brain age prediction performance across processing pipelines (raw, HARP, SIFT2, and HARP+SIFT2) for term and preterm groups. Metrics include mean absolute error (MAE, weeks), coefficient of determination (R^2^), Pearson correlation (r), and associated significance levels (p).

| Group | MAE (weeks) | R<sup>2</sup> | Correlation (r) | p |
| --- | --- | --- | --- | --- |
| **Raw** |  |  |  |  |
| Term | 0.758 | 0.525 | 0.728 | < 0.001 |
| Preterm | 1.060 | 0.438 | 0.662 | < 0.001 |
| **HARP** |  |  |  |  |
| Term | 0.757 | 0.525 | 0.728 | < 0.001 |
| Preterm | 1.057 | 0.440 | 0.664 | < 0.001 |
| **SIFT2** |  |  |  |  |
| Term | 0.787 | 0.470 | 0.686 | < 0.001 |
| Preterm | 1.061 | 0.442 | 0.665 | < 0.001 |
| **HARP+SIFT2** |  |  |  |  |
| Term | 0.781 | 0.476 | 0.692 | < 0.001 |
| Preterm | 0.995 | 0.481 | 0.695 | < 0.001 |

GA predictions for preterm infants across all connectomes resulted in higher prediction error compared to term infants. RAW and HARP connectomes showed similar performance, while SIFT2 weighting resulted in a minimal improvement in correlation. The HARP+SIFT2 pipeline showed a modest improvement in both correlation and MAE (Table 3).

Across all tractography pipelines, BAG estimates, reflecting the deviation between predicted and observed postmenstrual age, were centred close to zero in both term and preterm infants, indicating close alignment between predicted and observed age. In the term group (Figure 4), mean BAG ranged from −0.009 weeks for the raw connectomes to −0.016 weeks for the HARP+SIFT2 pipeline, while mean BAG in the preterm group ranged from −0.007 to −0.074 weeks. Thus, mean BAG estimates showed minimal variation between pipelines and were generally close to zero.

**Figure 4:**
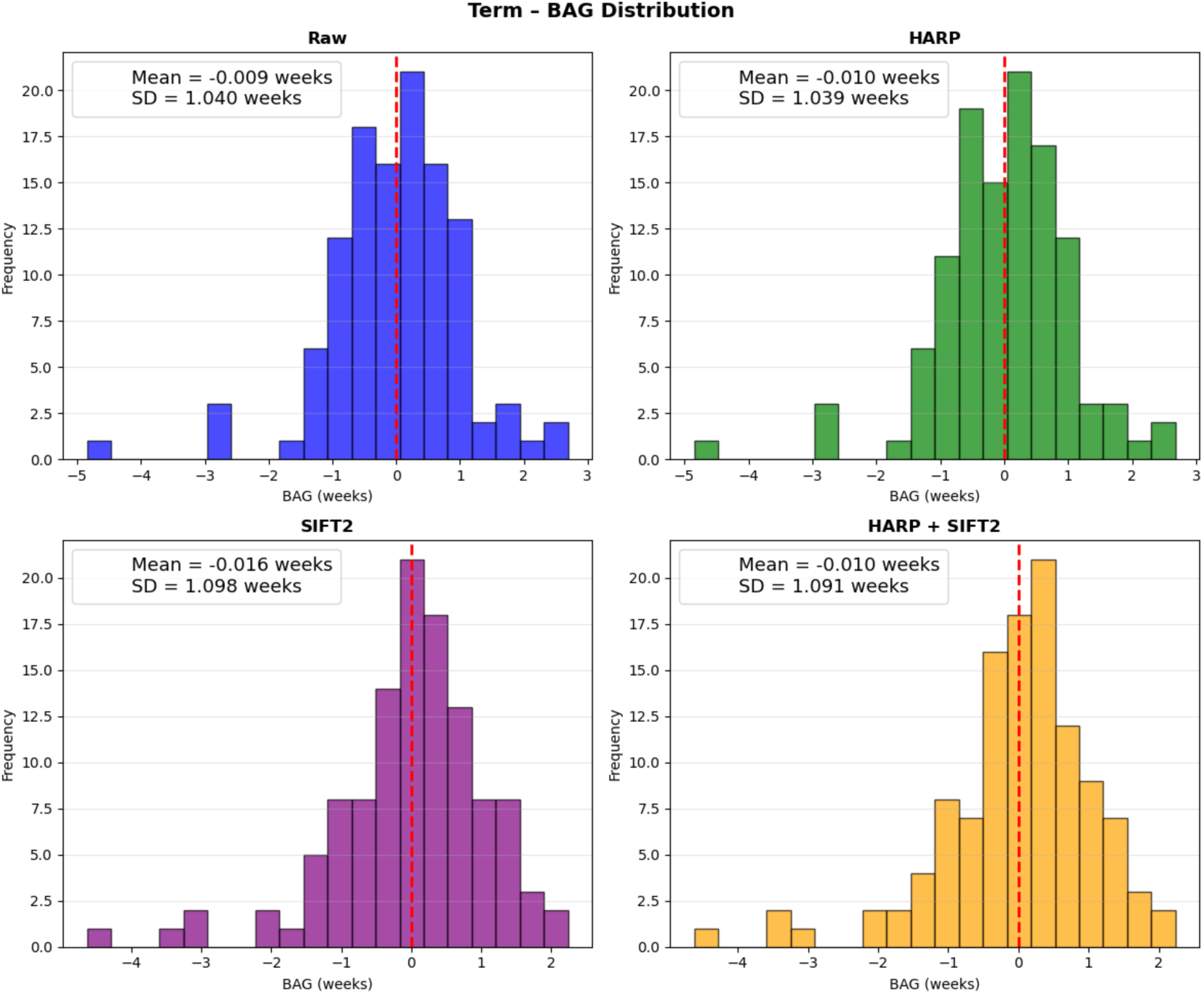
Brain age gap (BAG) distributions for term infants across processing pipelines (raw, HARP, SIFT2, and HARP+SIFT2). Histograms show the distribution of BAG estimates (weeks), with mean and standard deviation (SD) reported for each pipeline. The red dashed line indicates a BAG of zero, representing no difference between predicted and chronological age.

BAG variability was greater in preterm infants than in term infants across all pipelines. Standard deviations ranged from 1.383 to 1.442 weeks in the preterm group, compared with 1.039 to 1.098 weeks in the term group. Within the preterm group (Figure 5), variability was slightly reduced following filtering, from 1.442 weeks for the raw pipeline to 1.383 weeks for HARP+SIFT2. In the term group, SDs were relatively similar across pipelines, although slightly higher for SIFT2 and HARP+SIFT2 than for the raw and HARP pipelines.

**Figure 5:**
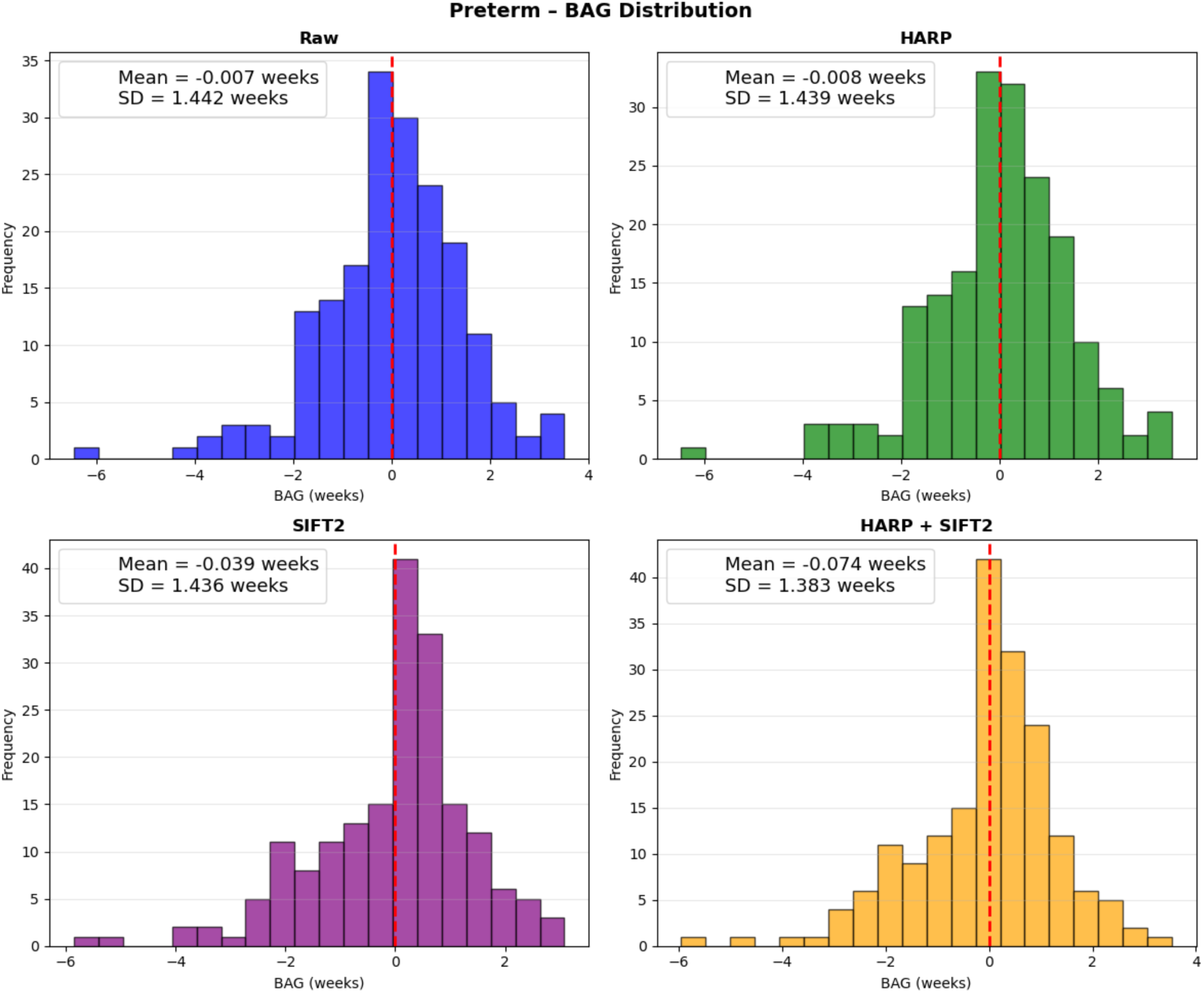
Brain age gap (BAG) distributions for preterm infants across processing pipelines (raw, HARP, SIFT2, and HARP+SIFT2). Histograms show the distribution of BAG estimates (weeks), with mean and standard deviation (SD) reported for each pipeline. The red dashed line indicates a BAG of zero, representing no difference between predicted and chronological age.

### 3.4. Connectome Analysis: Global Network Metrics

Global network metrics reflecting integration, segregation and hierarchical organisation of the structural connectome differed across the four processing pipeline configurations: raw, HARP, SIFT2, HARP+SIFT2 (Table 4). Significant differences between pipeline configurations were observed across all pairwise comparisons for global efficiency, transitivity, modularity, and complexity measures. The results demonstrated substantial differences in weighted network metrics between processing pipelines, although the pattern varied across metrics. Comparisons involving SIFT2 alone showed significant differences for most network measures, but did not significantly alter global efficiency relative to the raw pipeline. In contrast, the addition of HARP to SIFT2 resulted in a significant difference in global efficiency, indicating that the effects of tractography processing on network topology were dependent on the combination of processing steps. On the other hand, assortativity showed significant differences between raw vs. SIFT2 and between HARP vs. SIFT2, but not between comparisons involving the combined HARP+SIFT2 pipeline. These findings suggest that individual processing steps and their combination can differentially influence specific aspects of weighted network topology.

**Table 4.** Pairwise comparisons of weighted network metrics across pipelines for the full dataset (term and preterm). Connectivity matrices were normalised by the mean non-zero weight for each subject prior to calculation of network metrics, accounting for differences in connectivity weight between processing pipelines. P-values are reported for seven network metrics comparing raw, SIFT2, HARP, and HARP+SIFT2 pipelines.

| Network Metrics: Pipeline Comparisons |  |  |  |  |  |  |
| --- | --- | --- | --- | --- | --- | --- |
| Metric | Raw vs SIFT2 | Raw vs HARP | Raw vs HARP+SIFT2 | SIFT2 vs HARP | SIFT2 vs HARP+SIFT2 | HARP vs HARP+SIFT2 |
| Global Efficiency | 0.312 | **0.043** | **<0.001** | 0.318 | **<0.001** | **<0.001** |
| Average Local Efficiency | **<0.001** | **<0.001** | **<0.001** | **<0.001** | **<0.001** | **<0.001** |
| Transitivity | **<0.001** | **0.016** | **<0.001** | **<0.001** | **<0.001** | **<0.001** |
| Assortativity | **<0.001** | 0.115 | 0.288 | **<0.001** | **<0.001** | 0.294 |
| Modularity | **<0.001** | 0.289 | **<0.001** | **<0.001** | **0.022** | **<0.001** |
| Statistical Complexity | **<0.001** | **<0.001** | **<0.001** | **<0.001** | 0.735 | **<0.001** |
| Degree Variance | **<0.001** | **<0.001** | **<0.001** | **<0.001** | 0.544 | **<0.001** |
| **Note:** Significant p-values ($p < 0.05$) are shown in bold. | | | | | | |

Group-level differences between preterm and term infants across all pipeline configurations are presented in Table 5. Connectomes with both HARP+SIFT2 applied were the only configuration that showed significant differences (all *p* < 0.05) between the term and preterm groups across all network metrics (see HARP+SIFT2 column highlighted in Table 5). The distributions of network metrics for the HARP + SIFT2 connectomes are shown as violin plots in Figure 6, illustrating the distribution of metric values across the preterm and term groups. In general, term infants exhibited larger network metric values than preterm infants across most measures, as shown by the upward shift of the violin plots. This is consistent with the negative effect sizes observed in the HARP+SIFT2 configuration, reflecting greater values in the term group. Transitivity and statistical complexity deviated from this trend, showing lower values in term infants but higher values in the preterm group.

**Table 5.** Group-level comparison of network metrics between term and preterm infants across the four pipeline configurations. FDR-corrected p-values (bold when p < 0.05) and Cohen’s d effect sizes (positive effect size values denote higher values in the term group and negative values denote higher values in the preterm group) are reported for all network metrics.

| Network Metrics: Term vs Preterm |  |  |  |  |
| --- | --- | --- | --- | --- |
| Metric | Raw | SIFT2 | HARP | HARP+SIFT2 |
| Global Efficiency | **p < 0.001**<br>d = 1.389 | **p < 0.001**<br>d = -1.588 | **p < 0.001**<br>d = 1.386 | **p < 0.001**<br>d = -1.417 |
| Average Local Efficiency | **p < 0.001**<br>d = 0.943 | **p < 0.001**<br>d = -2.043 | **p < 0.001**<br>d = 0.938 | **p < 0.001**<br>d = -2.050 |
| Transitivity | **p < 0.001**<br>d = 0.540 | **p < 0.001**<br>d = 0.687 | **p < 0.001**<br>d = 0.540 | **p < 0.001**<br>d = 0.691 |
| Assortativity | **p = 0.022**<br>d = 0.267 | p = 0.119<br>d = -0.178 | **p = 0.023**<br>d = 0.266 | **p = 0.013**<br>d = -0.289 |
| Modularity | **p < 0.001**<br>d = -0.658 | **p < 0.001**<br>d = -0.385 | **p < 0.001**<br>d = -0.673 | **p < 0.001**<br>d = -0.411 |
| Statistical Complexity | p = 0.538<br>d = -0.072 | **p = 0.011**<br>d = 0.295 | p = 0.540<br>d = -0.072 | **p = 0.007**<br>d = 0.321 |
| Degree Variance | **p < 0.001**<br>d = 1.818 | **p < 0.001**<br>d = -0.897 | **p < 0.001**<br>d = 1.814 | **p < 0.001**<br>d = -1.002 |
| **Note:** Significant p-values ($p < 0.05$) are shown in bold. | | | | |

**Figure 6:**
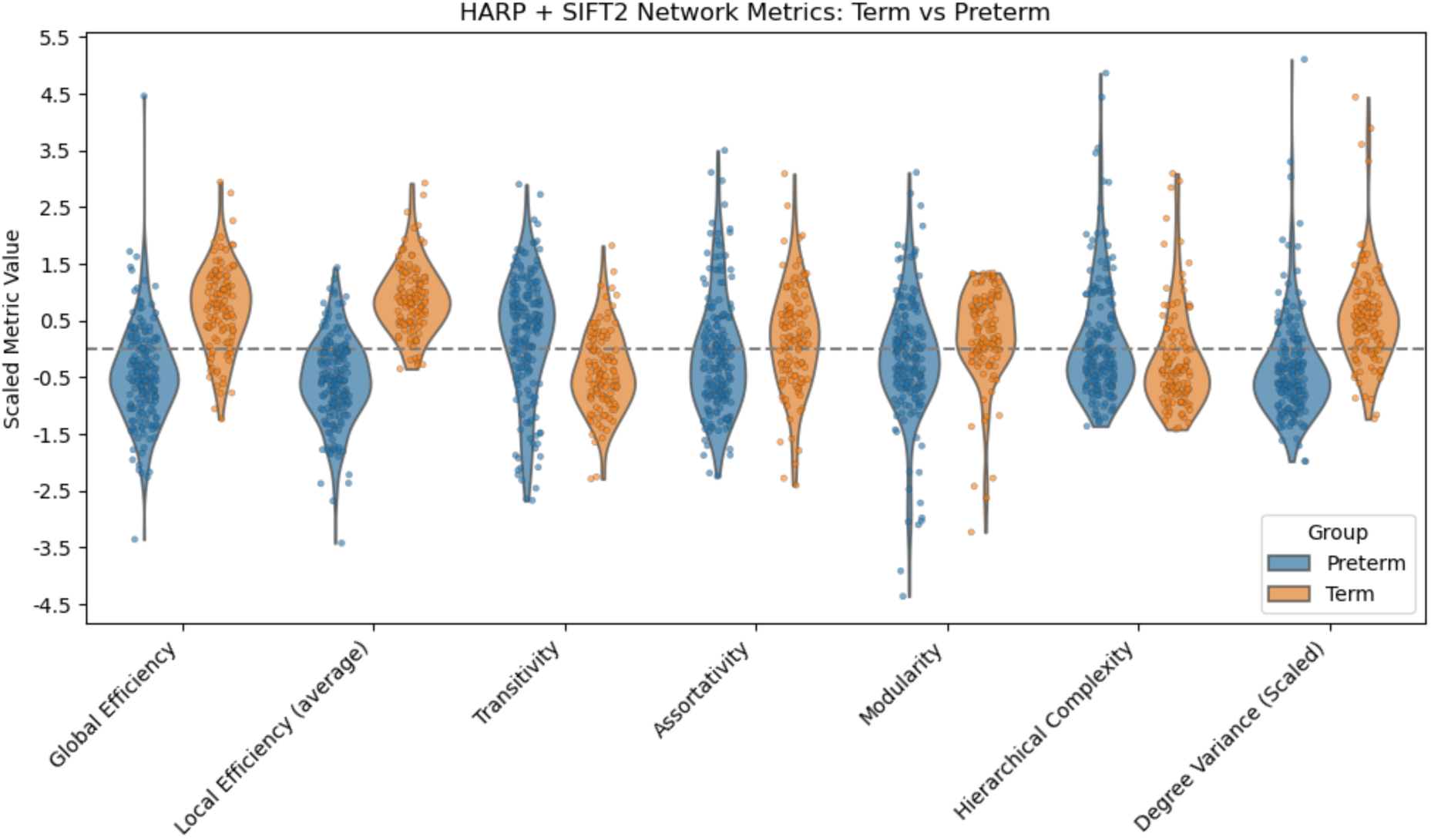
Comparison of term and preterm neonates across seven network metrics: Global Efficiency, Local Efficiency (average), Transitivity, Assortativity, Modularity, Statistical Complexity, and Degree Variance using the HARP + SIFT2 pipeline. Distributions are shown as z-score normalised violin plots to highlight group differences in structural connectivity.

## 4. Discussion

This study proposes a neonatal dMRI pipeline incorporating advanced tractography and post-processing techniques to support anatomically informed reconstruction of structural connectivity in early life, extending existing approaches to neonatal structural connectome construction. The proposed pipeline builds upon emerging advanced approaches to neonatal structural connectome reconstruction, including studies combining anatomically constrained tractography (ACT) with microstructure-informed streamline weighting, such as SIFT2 (Batalle et al., 2017; Blesa et al., 2019, 2021; Sa de Almeida et al., 2021; Taoudi-Benchekroun et al., 2022). We extend these approaches by implementing surface-based anatomical constraints and ensemble tractography, along with integrating HARP filtering with SIFT2 reweighting within an advanced neonatal connectome pipeline.

Brain age prediction was used as an independent validation, demonstrating that structural connectomes generated across all pipeline configurations produced robust predictions of GA in both term and preterm infants. Across all pipeline configurations, derived structural connectomes demonstrated the capacity to predict brain age in both term and preterm infants. MAE ranged from 0.76 to 0.79 weeks in term infants and 1.00 to 1.06 weeks in preterm infants, whilst correlations ranged from 0.48 to 0.53 and 0.66 to 0.70 respectively. Prediction accuracy was slightly higher in term infants, with MAE approximately 0.25 weeks lower than in preterm infants. BAG estimates were centred close to zero across all configurations, indicating minimal systematic bias, although BAG variability was greater in preterm infants than in term infants.

We then compared four tractography pipeline processing configurations (raw, SIFT2, HARP, and SIFT2+HARP) to examine how methodological choices influence global network topology across seven network metrics. Finally, term vs. preterm differences in metrics were evaluated separately for the four pipeline configurations to assess how tractography post-processing influences the detection of biologically meaningful group differences.

### 4.1. Validation using brain age predictions

Connectomes derived from all four pipelines were able to predict GA with good accuracy in both term and preterm infants. Brain age prediction performance is driven by two complementary aspects: MAE reflects absolute prediction accuracy in gestational weeks, whereas correlation reflects the ability of the model to preserve the relative ordering of brain maturation across subjects. The original study which presented the connectome-based predictive modelling (Sun et al., 2024) reported higher prediction errors in preterm compared with term infants. A similar pattern was observed in the present study, with prediction errors below one week in term infants and approximately one week in preterm infants (Table 3).

Moreover, the relative performance of the pipeline configurations differed between term and preterm infants. In term-born infants, raw and HARP produced the lowest prediction error (MAE = 0.76), while SIFT2 alone produced the highest MAE. Although these differences were small, combining HARP with SIFT2 improved prediction compared with SIFT2 alone (MAE = 0.78 vs. 0.79 weeks). Our results were comparable to those reported by Sun et al. (2024) which achieved an MAE of 0.92 weeks in term-born infants. Although prediction performance is likely influenced by differences in cohorts and prediction models between studies, comparisons with other existing work using neonatal structural connectomes for brain age prediction can provide a useful benchmark for evaluating the performance of our pipeline. The prediction performance of our connectomes was comparable to that reported in previous work predicting gestational age at scan in term-born infants, such as Taoudi-Benchekroun et al. (2022) who achieved MAEs of 0.84 and 0.72 weeks using random forest and deep neural network models, respectively.

In contrast, HARP+SIFT2 achieved the lowest prediction error in preterm infants, with an MAE below one week (Table 3), and showed the lowest variability in brain age gap (Figure 5). This pattern suggests that the combination of anatomical constraints and streamline reweighting may provide a more informative representation of structural brain maturation in preterm infants, where the structural organisation of white matter is more immature and variable. In comparison, the original study by Sun et al. (2024) reported an MAE of 1.80 weeks in preterm infants using the same model. Our results also represent an improvement over other reported brain age predictions from preterm structural connectomes. For example, Kawahara et al. (2017) reported an MAE of 2.17 weeks using a convolutional neural network, while Brown et al. (2017) reported an MAE of 1.55 weeks using a random forest regressor.

The improved performance of HARP+SIFT2 pipeline suggests that our combination of surface-based anatomical constraints, ensemble tractography, and complementary streamline filtering and reweighting produces connectomes that effectively capture biologically meaningful patterns of structural brain maturation. The results suggest that these advances improve the anatomical plausibility of reconstructed pathways while reducing the influence of implausible weighted streamlines, ultimately enhancing the developmental information captured by neonatal structural connectomes.

The BAG analysis provided further insight into these group differences. Mean BAG values were close to zero across all pipeline configurations in both term and preterm infants, suggesting little systematic bias in age prediction. However, BAG variability was consistently greater in the preterm group than in the term group, with higher standard deviations observed across all four configurations. The higher prediction error in preterm infants may reflect increased variability in brain development, including greater anatomical heterogeneity among preterm infants at term-equivalent age compared to term-born controls, as well as differences in GA at scan and its range between the groups. This may result in less consistent connectivity patterns, contributing to the higher MAE and BAG variability observed in the preterm group.

Overall, these results support the biological validity of the reconstructed connectomes and demonstrate their suitability for investigating developmental differences in neonatal brain networks. HARP+SIFT2 showed the lowest MAE for brain age prediction in preterm infants (1.00 week) and produced the lowest variability in BAG (SD = 1.38 weeks). However, differences in both prediction error and BAG variability across pipeline configurations were relatively small, suggesting that the choice of processing configuration had limited influence on overall brain age prediction performance. Moreover, the precision of the reference age should also be considered when interpreting differences between connectomes. Clinical guidance recognises that estimation of GA can be subject to inherent uncertainty (American College of Obstetricians and Gynecologists, 2017; Deepa et al., 2025), suggesting that the age used as the prediction target cannot be considered an exact ground truth for developmental brain age. Therefore, this inherent uncertainty warrants caution when interpreting differences in MAE as evidence that one pipeline meaningfully outperforms another in capturing developmental age, particularly when these differences are small. Predictive performance was broadly consistent across all connectome types, suggesting that structural connectivity patterns may retain meaningful information related to early brain maturation despite methodological difference. However, similarities in prediction performance may also suggest that brain age prediction is relatively insensitive differences in network organisation across pipeline configurations. Whilst brain age prediction may provide a useful global measure of developmental information within the connectomes, other metrics like graph theoretical measures can provide complementary insight into how processing choices influence the description of specific properties of neonatal brain network organisation.

### 4.2. Influence of post-processing on network topology

Pairwise comparisons of global network metrics derived from connectomes generated using four tractography post-processing configurations (raw, HARP, SIFT2, and HARP+SIFT2) revealed that methodological choices in structural connectome construction can influence the topology of neonatal brain networks. Connectomes using the SIFT2 pipelines differed significantly from raw and HARP tractograms across all global network metrics, highlighting the impact of post-hoc weighting on network organisation. Previous work has shown that tractography filtering techniques such as SIFT2 can alter structural connectivity estimates (He et al., 2024), but obtain more biologically meaningful estimates of structural connectivity (Frigo et al., 2020). Our findings extend this observation by demonstrating that methodological choices also influence global graph theoretical metrics derived from the resulting connectomes.

In contrast, the application of HARP alone resulted in relatively modest changes to global network metrics when compared with raw networks. Consistent with our findings, the original study also reported that HARP removed only a very small proportion of streamlines, with less than 1% classified as implausible (Leserri & Aydogan, 2024). However, when HARP was applied to clinical data in their most recent publication (Leserri et al., 2026), it removed almost 9% implausible streamlines. The discrepancy compared to our results could be explained from: (1) differences in pathway rules, (2) more rigid tracking parameters in our initial tractography (low curvature thresholds) and (3) less well-defined FODs from neonatal data. Despite this, the use of HARP is still argued to be an important step in the refinement of network reconstructions as it improves the overall anatomical plausibility of reconstructed streamlines.

These results suggest that different post-processing methods influence structural connectome topology in several ways. While SIFT2 substantially alters global network metrics through streamline re-weighting, HARP primarily acts as an anatomical refinement step by removing implausible streamlines without significantly altering large scale network organisation. Therefore, the combination of HARP and SIFT2 can redistribute weights to correct biases inherent to tractography and improve biological plausibility. In our data, only a small proportion of streamlines were removed, indicating that its effect is subtle yet meaningful, focusing on false positive connections involving implausible loops that do not reflect true white matter anatomy. Although differences between pipelines did not reach statistical significance for all metrics, the significant differences that were observed demonstrate that post-processing can influence derived network properties. Consequently, it suggests that network metrics may not be comparable between studies using different post-processing methods and should be interpreted with caution.

### 4.3. Influence of post-processing on detecting term vs. preterm group differences

When observing term vs. preterm differences between each pipeline configuration, all four connectome types identified significant group differences in the majority of network metrics. raw and HARP connectomes did not identify differences in statistical complexity, while SIFT2 connectomes did not detect differences in assortativity. However, the combination of HARP + SIFT2 was the only configuration that identified significant differences across every single global metric between the term and preterm groups. The ability of SIFT2 to improve sensitivity to group-level effects has been reported previously, with studies demonstrating enhanced detection of group differences and stronger associations with clinical measures following streamline reweighting (Frigo et al., 2020; McColgan et al., 2018). Such observations reinforce that SIFT2 modifies network topology and consequently influences network-derived metrics. In this study, the inclusion of HARP in tandem with SIFT2 presented the most comprehensive identification of term vs. preterm differences. This suggests that incorporating HARP alongside SIFT2 may reduce spurious information in network metric estimation, potentially improving the detection of group differences.

Examination of individual metric differences further highlighted the influence of tractography post-processing on biological interpretation. Within the HARP+SIFT2 pipeline, preterm infants exhibited lower global and local efficiency than term-born neonates, whereas raw and HARP connectomes suggested the opposite pattern, with higher efficiency observed in preterm infants. This sensitivity of network metrics to methodological choices may contribute to the inconsistent findings reported in the preterm connectomics literature (Zheng et al., 2023). While the present work was not designed to resolve these inconsistencies, our findings demonstrate that pipeline configurations have a profound effect both the magnitude and direction of observed group effects.

## 5. Conclusion

In this study, we proposed a neonatal structural connectome pipeline that builds upon existing approaches by integrating several methodological developments within a single framework. While recent neonatal connectomic studies have increasingly explored approaches beyond conventional DTI-based tractography, we attempt to contribute to this advancement by integrating anatomical surface constraints, ensemble tractography, streamline filtering with HARP, and streamline reweighting with SIFT2 within our pipeline. The resulting connectomes were used to test brain age prediction performance across the four pipeline configurations (raw, HARP, SIFT2, and HARP+SIFT2). Notably, the HARP+SIFT2 connectomes achieved a preterm brain age prediction MAE of less than one week which compares favourably with previously reported MAE of up to two weeks in other neonatal studies using structural connectomes. However, prediction performance was consistent across pipelines, suggesting that brain age prediction may be relatively insensitive to more subtle differences in network organisation introduced by the processing approach. Graph-theoretical measures therefore provide a complementary means of examining potential pipeline dependent differences in network organisation, and should therefore, be carefully considered in connectomic studies. We identified that groupwise differences between preterm and term-born neonates were most consistently detected in the HARP+SIFT2 connectomes. This finding suggests that combining anatomical refinement using HARP with streamline reweighting with SIFT2 may improve the sensitivity of connectomic analyses to subtle developmental differences. Most importantly, using HARP + SIFT2 derives the most biologically plausible connectomes, providing a more anatomically informed representation of structural connectivity while better reflecting the contribution of individual streamlines.

Overall, this study demonstrates that methodological choices in connectome construction can meaningfully influence analysis of brain network organisation, particularly in the context of preterm birth, where subtle differences in network topology may be sensitive to pipeline-dependent effects. For future work, evaluating the pipeline across multiple imaging centres and acquisition protocols would provide additional value in testing for robustness and generalisability. As a further avenue for exploring the utility of neonatal connectomes, brain age prediction could also be modelled using graph theoretical measures as model inputs, rather than the connectivity features themselves. Although beyond the scope of this study, implementing and testing this new model could provide insight into whether network-level measures provide comparable or additional predictive information to connectivity-based models.

## Data and Code Availability

Data included in this study is available to researchers under the terms of the Theirworld Edinburgh Birth Cohort Data Access and Collaboration policy available at https://reproductive-health.ed.ac.uk/theirworld-edinburgh-birth-cohort-tebc/for-researchers/data-access-and-collaboration.

The code used for the processing pipeline and scripts for network analysis can be found at: https://github.com/kwunh/neonatal_pipeline. The neonatal fork of trekker can be found at: https://github.com/dmritrekker/trekker_neonatal

## Author Contributions

Calvin Kai Hew: Conceptualization, Methodology, Software, Formal analysis, Data Curation, Writing - Original Draft, and Visualization; Simona Leserri: Methodlogy, Software and Writing - Review & Editing; Dogu Baran Aydogan Methodlogy, Software and Writing - Review & Editing; Mark E. Bastin: Data Curation and Writing - Review & Editing; Alexandra F. Bonthrone: Writing - Review & Editing; Kirsty Paterson: Investigation and Data Curation, Gayle Barclay: Data Curation, Charlotte Jardine: Data Curation, Keith M. Smith: Methodology and Writing - Review & Editing; Michael J. Thrippleton: Data Curation; Riccardo Marioni: Review & Editing and Supervision; James P. Boardman: Writing - Review & Editing, Supervision and Funding acquisition; Manuel Blesa Cabez: Conceptualization, Methodology, Resources, Data Curation and Writing - Review & Editing, Supervision.

## Funding

This work was supported by Theirworld (www.theirworld.org) and the Medical Research Council ([MR/X003434/1](https://www.sciencedirect.com/science/article/pii/S1878929326000563?via%3Dihub#spnsr2)). Participants were scanned in the University of Edinburgh Imaging Research MRI Facility at the Royal Infirmary of Edinburgh, which was established with funding from The Wellcome Trust, Dunhill Medical Trust, Edinburgh and Lothians Research Foundation, Theirworld, The Muir Maxwell Trust and other sources. DBA received funding from the Research Council of Finland (grant agreements #348631 and #353798), and Technology Industries of Finland Centennial Foundation (grant #4302). MBC is funded by a UKRI MRC programme grant (MR/X003434/1) awarded to Professor JPB, University of Edinburgh. CKH’s PhD studentship is funded by the Albert McKern Bequest, with additional funding from SINAPSE (Scottish Imaging Network: A Platform for Scientific Excellence) through the SECRE Award, which supported the collaboration between the University of Edinburgh and the CNIR group. We are grateful to the families who consented to take part in the study and to the radiographers at the Edinburgh Imaging Facility at the Royal Infirmary of Edinburgh for infant scanning.

## Declaration of Competing Interests

The authors have no conflict of interests.

## Appendices

### Appendix 1: Network Metrics Definitions

#### Global and Local Efficiency

Efficiency reflects how effectively information can be exchanged across a network. In the concept of brain connectivity, anatomical networks are considered to balance the opposing demands of functional integration and segregation (Rubinov & Sporns, 2010).

Global efficiency quantifies the overall integration of a network by measuring how efficiently information is exchanged across the entire system. It is defined as the average inverse of the shortest path length between all pair of nodes (Latora & Marchiori, 2001), capturing the efficiency of information transfer between brain regions within the network.

Local efficiency quantifies the extent of network segregation by measuring how efficiently information is exchanged within a node’s immediate neighbourhood (cluster) (Latora & Marchiori, 2001). It is also calculated using the average inverse shortest path length between all pairs of its neighbouring nodes, normalised by the total number of possible neighbour-to-neighbour connections.

#### Transitivity

Transitivity is another global measure of network segregation, but it is based on the prevalence of triangular connections within the network. It is calculated using the ratio of the total number of closed triplets compared to the total number of possible connected triplets (Newman, 2003).

#### Modularity

Modularity is a measure of network segregation that quantifies the extent to which a network can be partitioned into distinct communities (Newman, 2006). It quantifies the extent to which within module connectivity exceeds that expected under a random connectivity model. For each module, the observed connectivity is compared to its expected value, and this difference is summed across the network.

#### Degree assortativity

Assortativity is a measure of network resilience that quantifies the tendency of nodes to connect to other nodes with similar degree (Leung & Chau, 2007). Positive assortativity indicates a more cohesive network structure, in which high degree nodes tend to link with other high degree nodes, whereas negative assortativity suggests that high degree nodes connect mostly to low degree nodes, resulting in a more heterogeneous structure (Noldus & Van Mieghem, 2015). In brain networks, these properties may reflects the robustness of communication following disruption of highly connected hubs.

#### Degree variance

Degree variance is a measure of network heterogeneity, capturing the extent to which the network connectivity is dominated by highly connected hub regions (Smith & Escudero, 2020). Higher degree variance values reflect a more heterogeneous degree distribution, consistent with a hub rich network architecture.

#### Statistical complexity

Hierarchical relative strength variability (hRSV) is a weighted measure of statistical complexity that quantifies the diversity of connectivity patterns within a network (Yeung et al., 2025). The measure extends hierarchical complexity (HC), originally defined for binary networks using variability in neighbourhood degree sequences among nodes of the same degree (Smith et al., 2019). To accommodate weighted networks, hRSV computes the variability in relative node strengths among nodes with similar strength using a sliding window approach along the node strength hierarchy (Yeung et al., 2025). Higher hRSV values indicate greater diversity in connectivity organisation, whereas lower values are associated with more regular or random connectivity patterns.

## Notes

### Competing Interest Statement

The authors have declared no competing interest.

### Author Declarations

The study has ethics approval from the South-East Scotland Research Ethics Committee 02 (16/SS/0154 and 23/SS/0067) and NHS Lothian Research and Development (16/SS/0154 and 2023/0150).

